# Surgical Procedure Recognition Using Quantum Machine Learning

**DOI:** 10.1101/2025.08.21.25334146

**Authors:** Abdul Razak Nuhu, Peter Nimbe, Adamu Mohammed Mustapha, Eliezer Ofori Odei-Lartey

## Abstract

Surgical procedure recognition is the process of identifying tasks and gestures done during a surgical process and is a field that has been widely researched due to its use in robot assisted surgeries to improve surgical performances and training. This work investigates the use of Quantum Machine Learning (QML) algorithms, in particular Quantum Support Vector Classifier (QSVC), for the identification of patterns in kinematic data collected from the JIGSAWS dataset which includes 76 kinematic features related to suturing, knot tying and needle passing. In order to evaluate QSVC performance, we compared its performance measures such as accuracy, precision, recall and F1-score with that of a classical Support Vector Classifier (SVC). Quantum kernel-based methods like QSVM embed classical data into high-dimensional Hilbert spaces via quantum feature maps, offering the potential to capture complex data relationships more efficiently. Using ZFeatureMap and quantum circuits implemented in Qiskit, we demonstrate that QSVM shows slight performance advantages over its classical counterpart in specific tasks. These findings lay the groundwork for a context-aware surgical system to support medical practitioners in real time and help advance surgical practice and educational approaches for enhancing patients’ quality of life.

## Introduction

General surgery is the core of surgical practice and involves the whole scope of surgical procedures for diagnosing, treating, and managing disease conditions across different organs with multiple systems(Kulkarni). From emergency interventions to elective procedures, general surgeons’ training enables them to handle a wide range of surgical issues with accuracy, competence, and kindness(Jain & Puranik, 2022). Initially, general surgery has had a lot of developments in the past decades and this is due the increased knowledge in medical field, surgical techniques, and technology (Farivar, Flannagan, & Leitman, 2015). Innovations like anesthesia, aseptic techniques, and minimally invasive techniques have changed the whole surgery field and as a result, it makes surgeries safer, more effective, and less invasive for the patients (Riskin, Longaker, Gertner, & Krummel, 2006). In the recent era, general surgery is still at the leading edge of the advances in medical science and innovation, where the main concern is the personalized, evidence-based medicine and multidisciplinary collaboration. Modern surgical approaches are interested in exploiting the most advanced tools to cut surgical errors and improve the patient experience (Reddy et al., 2023). Quantum machine appears to be a topical tool of interest.

Quantum machine learning and the general surgery represent a groundbreaking development in surgical science, where quantum algorithms help to process complex medical data, predict results, and improve procedural workflows(Tarassoli, 2019). Quantum machine learning based systems combined with the surgical environment provide real-time information, adaptive guidance, and tailored recommendations to surgeons improving them to take decisions, have precision, and efficiency in the operating room (Solenov, Brieler, & Scherrer, 2018). The algorithms of QML can be in continuous learning and evolution, taking from the huge datasets of surgical experiences to adjust and improve their predictive models and the patients care pathways. Surgical robots with this capability of adaptive learning may open doors for progressing surgical techniques, bettering patient outcomes, and observing the transformation of general surgery(Qian & Ren, 2025). While surgeons face the challenges of contemporary healthcare, they still remain dedicated to the goal of providing excellent care for the patient which is centered on the patient with the use of innovative technologies and ideas. One aspect of these challenges lies in the effectiveness and efficiency of wound closure during surgery.

Needle passing, suture application, and knot tying are the basic operations of wound closure. Entailing specific steps such as reaching for and positioning the need, approximation of tissue before pushing the needle, and orienting the needle and moving from beginning points to end points. The quality of wound closure has significant influence on the recovery of the patient (Khan, Bann, Darzi, & Butler, 2003). Inefficient wound closure can lead to surgical complexities including wound dehiscence, tissue damage, or surgical site infections, which may prolong hospital stays, raise the cost of healthcare, and undermine the quality of care (Jackson, Desai, Castillo, & Çavuşoğlu, 2016). Surgery operations can be fast paces, and so even with experts, the effectiveness of these operations can be constrained by short time windows in the operating room, mounting pressure on surgeons to pace faster and increasing the chances of errors during the needle tying or suturing process (Mohan & Dalal, 2013).

Wound closure operations require extensive training and experience to be precise. Mastering of such skills is a gradual process, particularly for beginners, who might end up causing harm to several patients during the learning process (Sarker & Vincent, 2005). Current process for managing errors during wound closure are manually driven by experts, who observe progress and give subjective feedback on quality of work. However, the manual method is tedious and easily compromises the quality of progress monitoring and feedback. In addition, there are no objective metrics for measuring quality and so providing standardized training for beginners cannot be guaranteed (Fard, Ameri, & Ellis, 2016). Advance technologies like robotic assistance are available to improve the quality of wound closure. However, very limited budgets are available for this purpose (Elgezua, Kobayashi, & Fujie, 2013). Resolving these challenges will be a product of creative process which will use the most recent technologies like artificial intelligence and machine learning systems (Nasser, 2018) that may not require extensive cost on additional infrastructure.

The quantum support vector machine (QSVM) implements kernel-based approach through utilization of parameterized quantum circuits for converting classical data into quantum states. The constructed feature map through these circuits enables QSVMs to solve difficult classification problems by mapping inputs into high-dimensional Hilbert spaces. Quantum circuits outperform classical machines because they can execute complex kernel functions which classical machines cannot process. The potential of quantum systems makes them suitable for intricate surgery gesture recognition when fine details in data need to be distinguished.

## Related Work

Surgical procedure recognition is the process of determining specific tasks and gestures that are accomplished during surgery. This field has attracted much interest because of the possibility of improving the outcomes of surgical operations through automation and the use of robots. The assessment of the specific tasks involved in surgery like suturing, needle passing, and knot tying is important for creating an intelligent surgical system that can help the surgeon in real-time, give feedbacks, and enhance the training results (Eckhoff et al., 2023). Classical computing has greatly enhanced the area of suturing in surgical healthcare through the enhancement of training models, robotic surgery, and feedback mechanisms (Shen et al., 2018). The use of virtual reality (VR) and augmented reality (AR) in surgical training has revolutionized the manner in which suturing skills are honed through the use of simulation. These systems help the surgeons to practice suturing in a safe environment, with high risk of error, but with the opportunity to practice as much as possible and get feedback right away (Shen et al., 2018). A review by (McKnight et al., 2020) shows how the use of VR and AR simulations has improved the training of surgeons especially in the mastery of suturing through practice and feedback. Quantum computing is a key to the sphere of medicine and is able to solve a great number of old problems which have never found solution till now. It can help to advance diagnostics, treatment optimization, drug discovery and individualized medicine (Hassanzadeh, 2020). Quantum algorithms can provide fast and reliable way of healthcare delivery system improvement by evaluating a large-scale healthcare data, finding inefficiencies and optimizing resource allocation(Tarassoli, 2019). The role of quantum computing can be seen in the scheduling of patient appointments, allocation of resources, and prediction of disease outbreaks(Srikanth & Kumar, 2022).

Quantum algorithms are capable of simulating molecular interactions and can predict drug efficacy with an unmatched precision that brings the discovery of novel therapeutic compounds into the sprinter zone and helps to optimize the existing drugs as well(Maniscalco et al., 2022). Enormous potential of quantum computing is clear for the transformation of the healthcare industry environment, bringing infinite power of computation to the most difficult issues of the healthcare sector(Flöther, 2023). The quantum computing applications are targeted to automate the complicated processes of healthcare providers, and so, by the means of quantum algorithms and technologies, healthcare providers can unlock new insights, accelerate medical innovation, and ultimately improve the quality of care for patients worldwide(Srikanth & Kumar, 2022).

Some of the classical machine learning models that have been used in the JIGSAWS dataset include SVM, Random Forest, and k-NN. However, recent studies have shown that deep learning approaches, such as convolutional neural networks, can outperform these traditional methods in terms of accuracy and generalization (Ismail Fawaz, Forestier, Weber, Idoumghar, & Muller, 2018). Recently, CNNs and RNNs have been used to recognize the surgical procedures due to their effectiveness in deep learning. These models can learn the features from the raw data in an unsupervised manner which saves the time of feature extraction(Huo et al., 2017). For instance, CNNs can help analyze video frames to identify the visual features of various surgical tasks, while RNNs can capture temporal relations in the kinematic data(Hamdi, Aboeleneen, & Shaban, 2021).

Researches on the JIGSAWS dataset have shown that deep learning models are more accurate and less sensitive to noise compared to the classical models (Qin et al., 2020). To the best of the authors’ knowledge, there are only a few studies exploring QML for surgical applications, but the results indicate that quantum models could process the high-dimensional data created during surgeries effectively (Mathur et al., 2021), leading to improved classification accuracy compared to classical deep learning models, especially when the training datasets are small (Matic, Monnet, Lorenz, Schachtner, & Messerer, 2022). However, there are many challenges that arise in the practical application of QML such as the requirement of quantum computing hardware and the creation of quantum algorithms for the specific data of surgery (Cerezo, Verdon, Huang, Cincio, & Coles, 2022). Hybrid quantum-classical models present a promising middle ground, where some operations are performed on a quantum device and the rest on classical hardware, thus mitigating the hardware requirements (De Maio et al., 2024). QSVM is a quantum variant of the classical SVM which is a supervised machine learning algorithm used for classification and regression. QSVM uses the concepts of quantum computing to possibly offer benefits in solving classification issues especially when the classical SVMs encounter computational issues(Innan, Khan, Panda, & Bennai, 2023).

The table 1 below presents related works that has been done using classical machine learning models

**Table 1:**
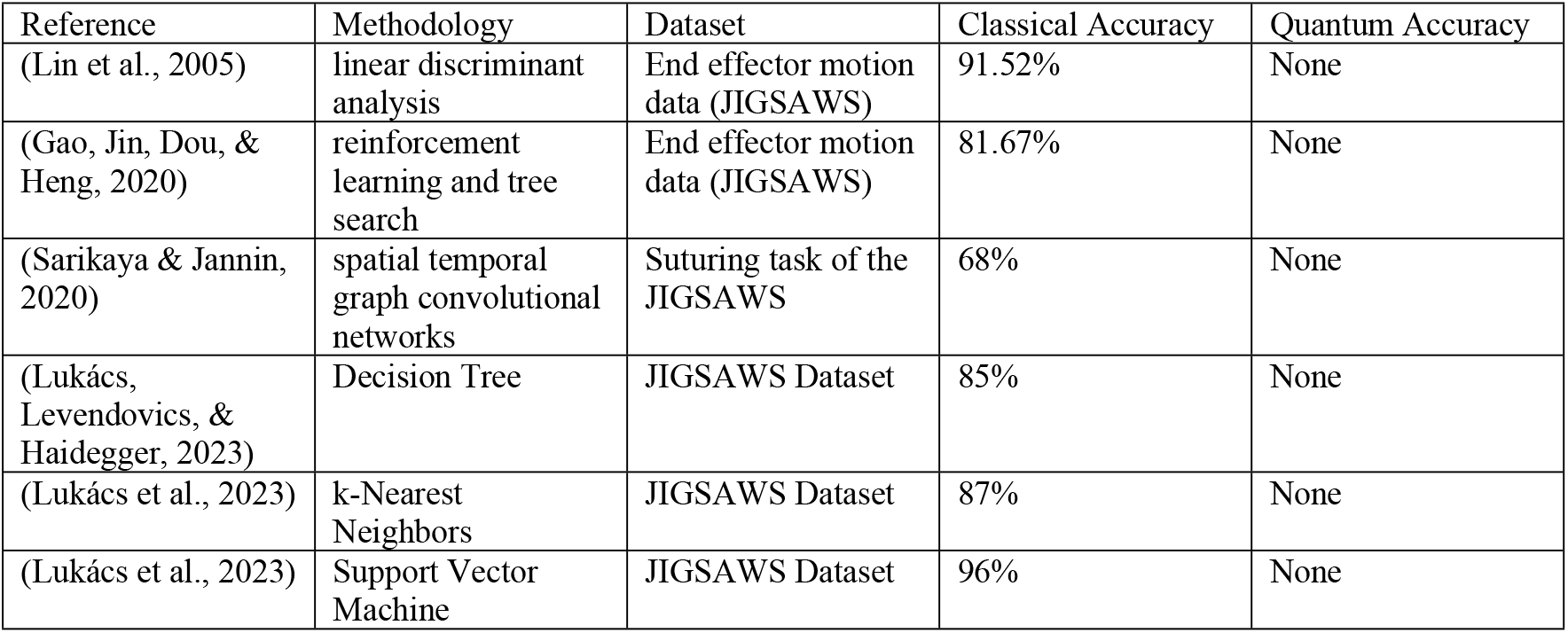
Related works.

These results indicate that there has been little work in the application of quantum machine learning in surgical healthcare which is an area with great potential to help improve surgical healthcare using machine learning to improve patient outcomes.

## Methods

### Data Acquisition

The kinematic data used for this study was obtained from the JIGSAWS (JHU-ISI Gesture and Skill Assessment Working Set) dataset, which includes measurements related to the movement of surgical tools during tasks such as suturing, knot tying, and needle passing. The dataset comprises 76 kinematic features per time frame, including tool positions, velocities, rotations, and gripper angles.

The dataset contains 76 kinematic features per time frame, such as:

- **Tool Positions (X, Y, Z coordinates)**
- **Tool Velocities** (translational and rotational)
- **Tool Rotations** (roll, pitch, yaw)
- **Gripper Angle**

### Data preprocessing

This is essential for both classical and quantum models to provide high quality and relevant data. The following preprocessing steps are applied:

### Data Cleaning

Missing values are addressed by either of the following: deletion of samples with missing values or by filling the missing values.

### Normalization

The kinematic data is normalized in a manner that is common to all features such as the Min-Max scaling. This step is critical for models based on SVM, because these models are very sensitive to the scaling of features.

### Label Encoding

The surgical tasks namely suturing, knot tying, and needle passing were represented by numerical tags (0, 1, 2) for classification by the machine learning models.

### Classical Model Development

In the case of the classical machine learning model, a Support Vector Machine (SVM) classifier was used. SVM is an effective algorithm in classification problems particularly when analyzing high dimensional data which is the case with kinematic data in this study.

#### Model Configuration

The classical SVM is set up with a RBF kernel that is appropriate for nonlinear classification problems. To optimize the C and gamma parameters, grid search and cross-validation methods are used.

#### Training

SVM model is applied on kinematic data after preprocessing, with the features being the tool movements and the labels being the surgical task (suturing, knot tying, or needle passing).

#### Performance Metrics

The model is assessed by using accuracy, precision, recall, F1-score and confusion matrix to guarantee an overall assessment of the classification model.

### Quantum Model Development

Quantum Support Vector Machine (QSVM) was implemented using Qiskit machine learning model. QSVM uses a quantum enhanced kernel function to map data into a high-dimensional Hilbert space, allowing the model to identify complex patterns within the surgical kinematic data.

### Quantum Feature Mapping and Embedding

The kinematic data was transformed into a higher-dimensional quantum space by applying a quantum feature map using ZFeatureMap from Qiskit which applies a series of parameterized rotations along the Z-axis of each qubit followed by entangling gates. This mapping encodes the classical data into quantum states which allows the quantum model to leverage quantum characteristics to model intricate connections found in the data. ZFeatureMap was selected because it preserves data locality while increasing feature expressivity, which is beneficial for high-dimensional datasets with limited samples. It also supports modular depth configuration through the **reps** parameter, which helps control the circuit complexity.

### Quantum Circuit

QSVM was constructed using Qiskit, where quantum circuits are used to implement the quantum feature map and decision plane. These circuits use quantum gates including Hadamard gates for superposition which to represent multiple input states simultaneously, CNOT gates for entanglement and rotation gates for phase and amplitude adjustments were used to process the kinematic data to exploit correlations between input features.

### Data Dimensionality Reduction

Prior to quantum embedding, Principal Component Analysis (PCA) was applied to reduce the original 76 features to a smaller subset suitable for quantum circuits. This step was necessary to manage the number of qubits required and to reduce noise in the data. This reduced feature set was used to define the number of qubits in the quantum circuit.

### Experimental Setup and Execution

The dataset was split into three surgical tasks: suturing, knot tying, and needle passing. Each was preprocessed independently. The features were normalized using MinMaxScaler and transformed using PCA (n=5). The resulting datasets were split into training (80%) and testing (20%) using train_test_split with a fixed seed of 42.

The QSVM classifier was configured with:

- **Backend**: Qiskit BasicAer statevector_simulator
- **Quantum kernel**: Fidelity-based kernel computed from overlaps of quantum states
- **Qubits**: 5 (equal to PCA components)
- **Feature Map**: ZFeatureMap with 2 repetitions
- **Regularization Parameter**: C=1000
- **Random Seed**: 12345 for reproducibility

Both classical and quantum models used the same training and test splits for fair comparison. Cross-validation was conducted using 5 folds.

### Reproducibility

The experiments were conducted using the following software stack:

- Python 3.9.2
- Qiskit 0.46.2
- Scikit-learn 1.5.1
- NumPy 2.0.2
- Matplotlib 3.9.2

All preprocessing, model training, and evaluations were done on a machine with Intel Core i5-1145G7 (4 cores, 8 threads) and 16GB RAM. The QSVM was simulated using the Qiskit Aer qasm_simulator.

For classical SVM, we used RBF kernel with hyperparameters tuned via grid search:

- C: [0.1, 1, 10]
- gamma: [‘scale’, 0.01, 0.001]

For QSVM:

- Feature map: ZFeatureMap (reps=2)
- Backend: AerSimulator
- Shots: 1024
- Dimensionality reduction: PCA with 5 components applied to kinematic features prior to embedding

Train-test splits were stratified and used consistently across classical and quantum pipelines to ensure a fair comparison. Random seeds were fixed using algorithm_globals.random_seed = 12345 for consistent simulation outcomes.

The complete code used for training and testing the models, including data preprocessing and cross-validation routines, is available upon request to support full reproducibility of results.

### Training

The QSVM is trained with the same kinematic data as the classical SVM which makes it possible to compare them. Several quantum optimization algorithms were employed to reduce the classification error, and the quantum kernel function playing a crucial role in data classes.

### Training and Testing Process

The performance of both the classical SVM and QSVM models was evaluated using the preprocessed kinematic data from the JIGSAWS dataset.

### Cross-Validation

To increase the generalization ability of both the models, k-fold cross-validation method is used. The data was divided into k sets, with the model trained om k-1 sets and tested on the remaining. The above process was iterated k times to achieve high accuracy in performance assessment.

### Hyperparameter Tuning

In both models, hyperparameter tuning was performed. For the classical SVM penalty term (C) and kernel width (gamma) were tuned using grid search and cross validation. In the case of QSVM, the quantum feature map and the depth of quantum circuit was adjusted to optimize the performance.

### Test Set Evaluation

After the training, both models were evaluated on a test set with the purpose of identifying how accurate the models are in predicting new data.

### Model Evaluation

The performance of both the classical SVM and QSVM models was evaluated using several metrics:

#### Accuracy

The total percentage of all the instances which have been classified correctly.

Precision, Recall, and F1-Score: Specificity is the ratio of true positive to the sum of true positive and false positive; sensitivity is the ratio of true positive to the sum of true positive and false negatives; F1-Score is the harmonic mean of specificity and sensitivity.

#### Confusion Matrix

A confusion matrix is used to identify how the models distinguish the classes (the surgical tasks). Quantum Advantage: The comparison also seeks to find instances where QSVM might be superior to classical SVM, referred to as quantum advantage.

## Results

The study entails the use of a large amount of computational power for model training and assessment. Classical models are trained using high-performance CPUs, whereas quantum models are trained using available quantum simulators and hardware.

We used a Dell Intel Core i5-1145G7 is a quad-core processor with a base clock of 2. 60 GHz and a max turbo frequency of 4. 40 GHz and 16gb RAM. It has a four-core eight-thread design, which allows for parallel processing and increases the efficiency of machine learning computations, quantum simulations, and classical models. Executing QSVM or any other quantum algorithms with the help of Aer simulator from Qiskit consumes a lot of computational power, especially when working with large quantum circuits. The 16 GB of RAM assists in this, but as quantum circuits become larger, memory is an issue. For the classical machine learning tasks (e. g. Random Forest, SVM, QSVM kernel methods) there is enough computational power in the Intel i5 processor with 8 logical cores to perform parallel computations like cross validation.

The evaluation was done based on the accuracy, recall and F1-score for the gesture classes. The classical model, Support Vector Classifier (SVC), yielded a good accuracy for all the gesture classes with an accuracy of 96% as indicated in **table 2**. The precision, recall, and F1-scores were high for each of the gesture classes and all the scores were between to 0.91 and 1.00. This means that the model was able to recognize the differences in the various suturing gestures as it was able to classify them correctly. Likewise, the QSVC model had an accuracy of 99% which is quite impressive and has a good potential for the classification of the data. Precision, recall, and F1-scores of most gesture classes were almost 1.00, which proved that the quantum model was able to identify the intricate movements involved in the suturing task with the better level of success as the classical model for some classes but overall, the QSVC performed better in accuracy.

**Table 2.** Result for suturing classification.

| Suturing Task Classification |  |  |  |  |  |  |
| --- | --- | --- | --- | --- | --- | --- |
| Class | SVC Precision | SVC Recall | SVC F1-Score | QSVC Precision | QSVC Recall | QSVC F1-Score |
| Reaching for needle with right hand | 0.91 | 1 | 0.96 | 1 | 1 | 1 |
| Positioning needle | 0.94 | 0.91 | 0.92 | 0.99 | 0.97 | 0.98 |
| Pushing needle through tissue | 0.96 | 0.99 | 0.97 | 0.98 | 1 | 0.99 |
| Transferring needle from left to right | 1 | 0.99 | 0.99 | 1 | 0.98 | 0.99 |
| Moving to center with needle in grip | 1 | 0.80 | 0.89 | 1 | 0.97 | 0.99 |
| Pulling suture with left hand | 0.98 | 0.98 | 0.98 | 0.98 | 0.97 | 0.98 |
| Orienting needle | 0.96 | 0.95 | 0.95 | 0.99 | 0.98 | 0.99 |
| Dropping suture at end and moving to end points | 1 | 1 | 1 | 1 | 1 | 1 |
| **Accuracy: SVC (0.96), QSVC (0.99)** |  |  |  |  |  |  |
| **Macro Avg: SVC (Precision: 0.97, Recall: 0.95, F1: 0.96), QSVC (Precision: 0.99, Recall: 0.98, F1: 0.99)** |  |  |  |  |  |  |

The high correlation between the test accuracy and the cross-validation scores for both the support vector and quantum support vector in **figure 1** also show that the model is not underfitting because the performance of the model is almost the same for the different validation sets. The small difference in the cross-validation scores shows the ability of the model to perform on other sections of the data set. The training loss decreases as the number of training samples increases, this indicates a good learning for the model.

**Figure 1:**
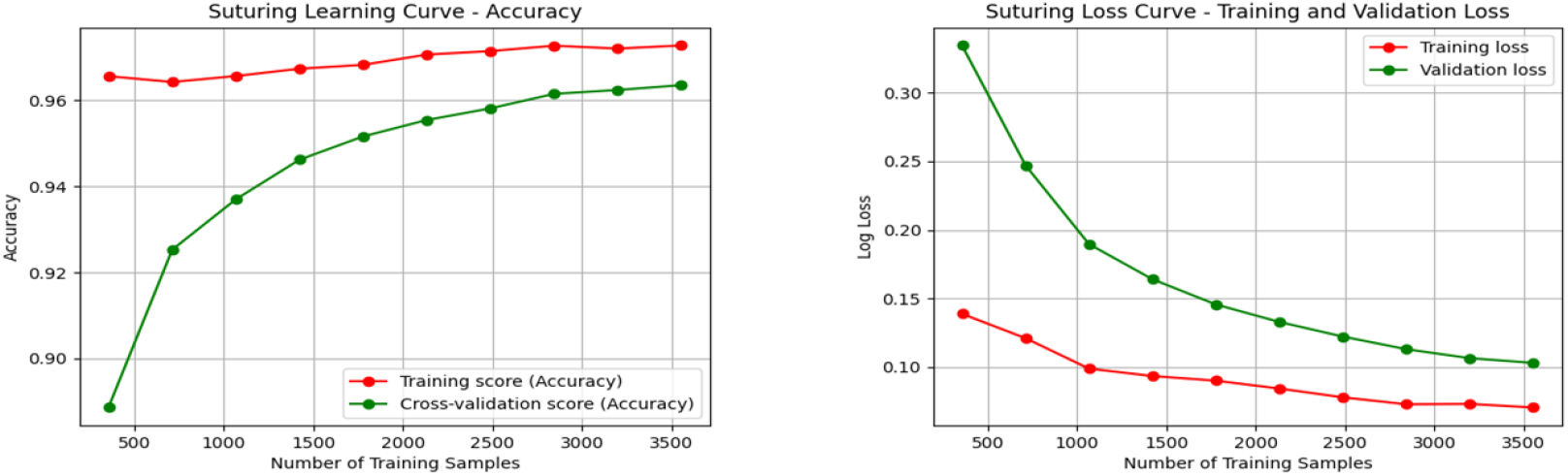
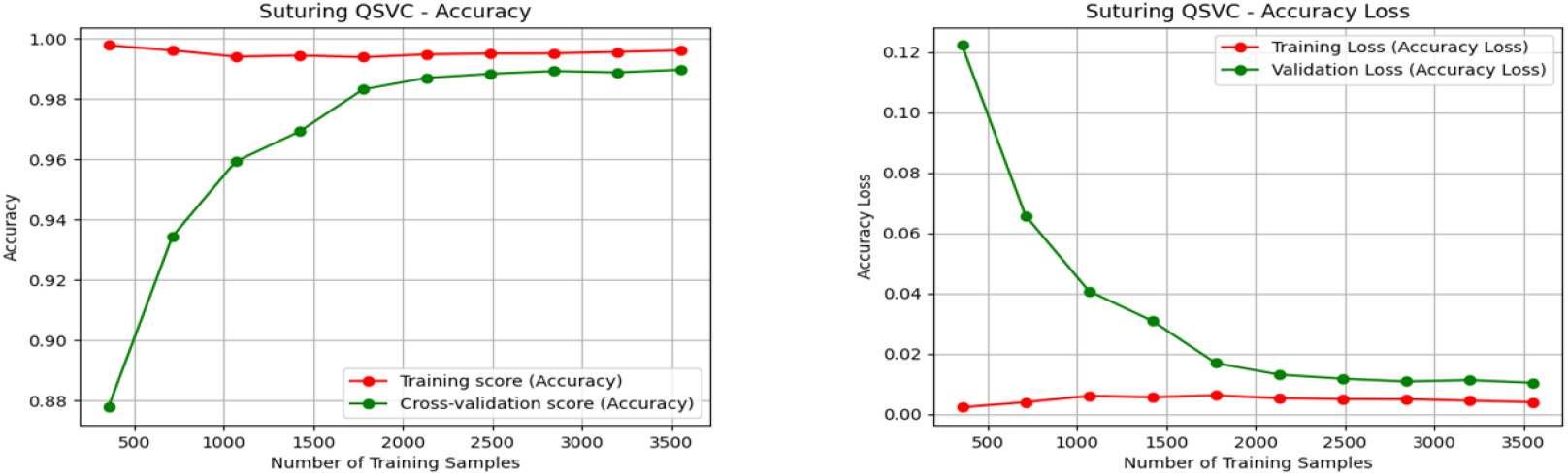
Suturing Training Accuracy and validation loss

In table 3, we compare the outcomes of classical algorithms, that is, Support Vector Classifier (SVC), and Quantum Support Vector Classifier (QSVC). The evaluation is done with reference to the precision, recall and the F1-scores of the gesture classes that have been identified.

**Table 3:**
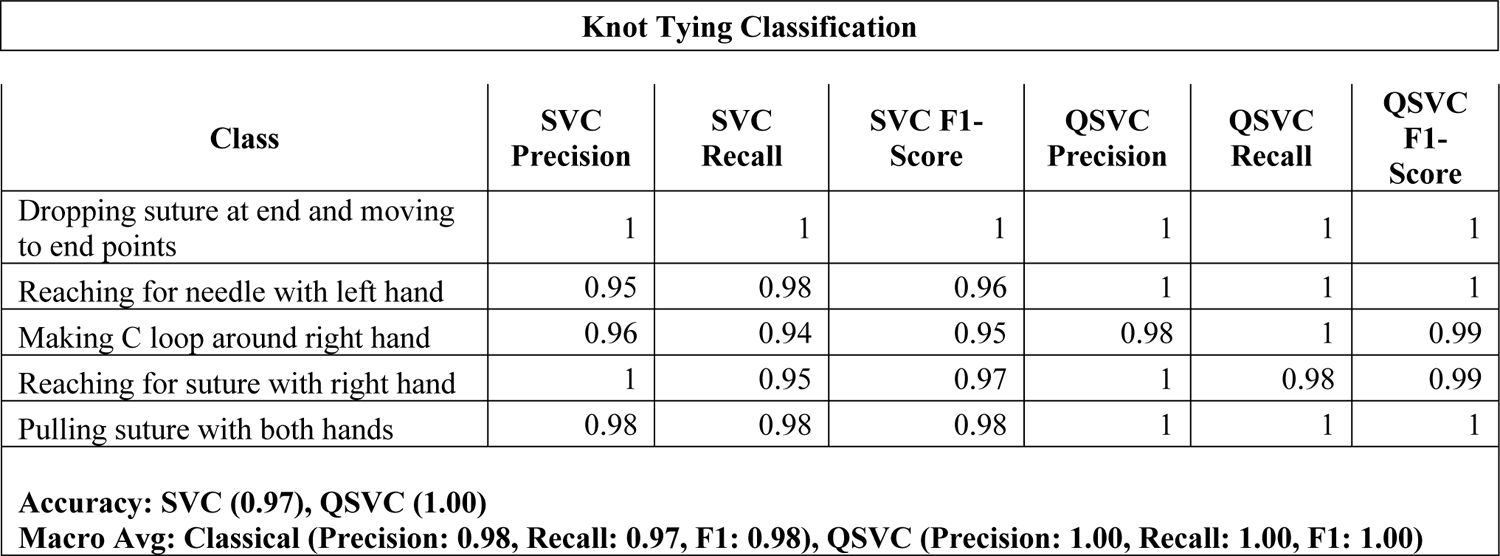
Knot tying classification.

| Knot Tying Classification |
| --- |

| Class | SVC Precision | SVC Recall | SVC F1-Score | QSVC Precision | QSVC Recall | QSVC F1-Score |
| --- | --- | --- | --- | --- | --- | --- |
| Dropping suture at end and moving to end points | 1 | 1 | 1 | 1 | 1 | 1 |
| Reaching for needle with left hand | 0.95 | 0.98 | 0.96 | 1 | 1 | 1 |
| Making C loop around right hand | 0.96 | 0.94 | 0.95 | 0.98 | 1 | 0.99 |
| Reaching for suture with right hand | 1 | 0.95 | 0.97 | 1 | 0.98 | 0.99 |
| Pulling suture with both hands | 0.98 | 0.98 | 0.98 | 1 | 1 | 1 |
| **Accuracy: SVC (0.97), QSVC (1.00)**<br>**Macro Avg: Classical (Precision: 0.98, Recall: 0.97, F1: 0.98), QSVC (Precision: 1.00, Recall: 1.00, F1: 1.00)** |  |  |  |  |  |  |

The classical model, Support Vector Classifier (SVC), was identified to be very efficient in the knot tying task with the overall accuracy of 97%. The precision, recall, and F1-score of all the gesture classes were high with some of the gestures having 100%. The model had a slightly lower precision for class of 0.95 for some classifications which may indicate the difficulty in discriminating this gesture from other gestures, the recall was 97% percent, while the quantum model (QSVC) was slightly better with accuracy of 100 percent compared to the classical model. The precision, recall and F1-scores of each of the gesture classes for QSVC were between 0.98 and 1.00 for all the classes as shown in the following table below. The minor variations in the accuracy of the classical model for some classes were not observed in the QSVC model which shows that the QSVC model is better in generalizing across the knot tying gestures.

In **figure 2**, the correlation between the test accuracy and the cross-validation scores for both the support vector and quantum support vector was very high but the QSVC seems to perform better as data increases as compared to the SVC though the SVC performs well as the data increases, this shows that the model is not underfitting because the performance of the model is almost the same for the different validation sets. The small difference in the cross-validation scores shows the ability of the model to perform on other sections of the data set. The decreasing validation loss shows that the model is generalizing better as it is exposed to more data.

**Figure 2:**
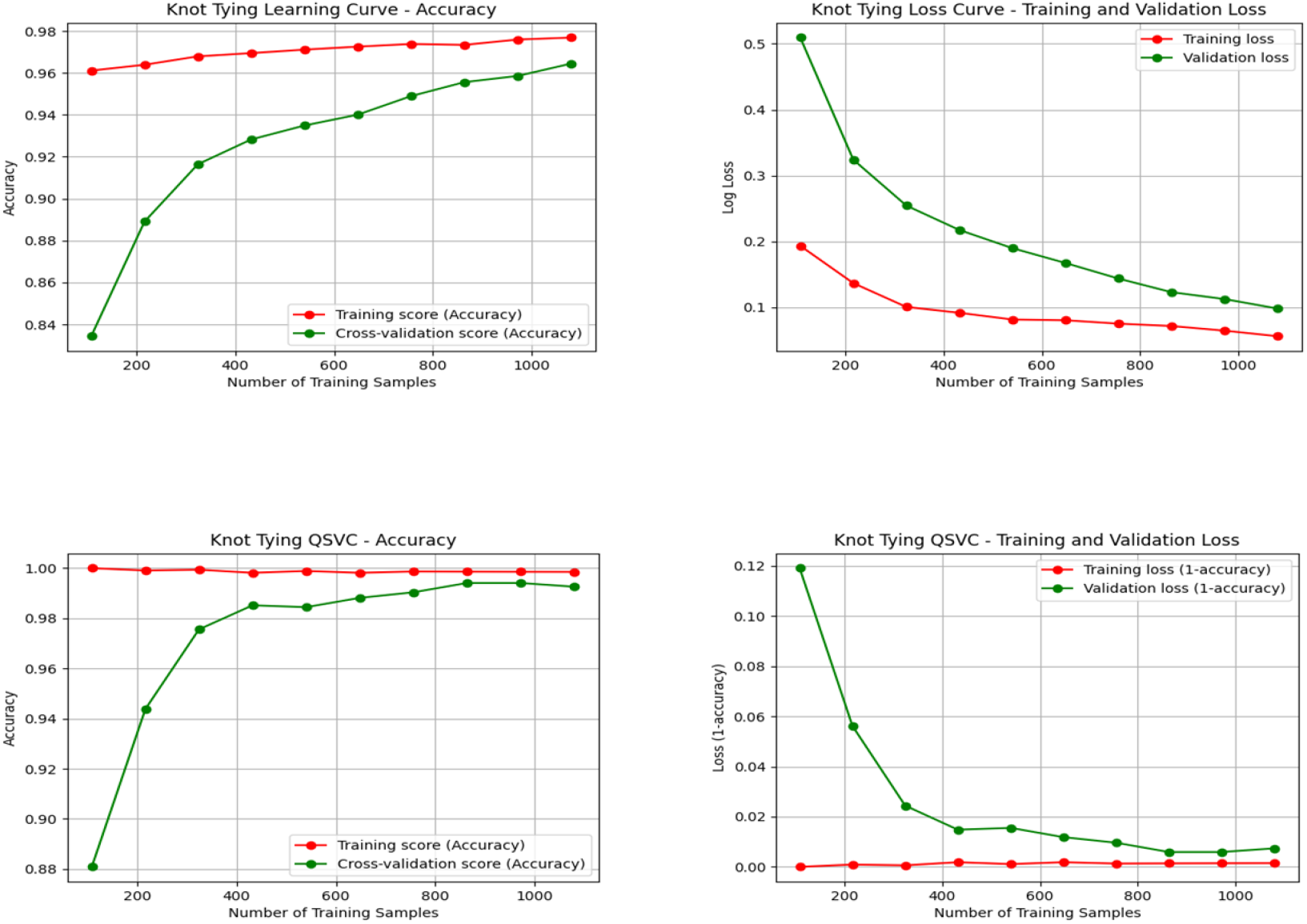
Knot tying Training Accuracy and validation loss

In table 4 below, the Classical Support Vector Classifier (SVC) model yielded an overall accuracy of 96% in the needle passing task with high accuracy in all the gesture classes. In most cases, the precision, recall, and F1-scores were 95% for all the gestures with a slight variation in recall for some classes. However, these differences are insignificant, and the overall model’s performance demonstrates its reliability in identifying needle passing gestures whereas the QSVC model was also found to have an overall accuracy of 99% which is higher than the classical model.

**Table 4:** Needle passing classification.

| Needle Passing Task |  |  |  |  |  |  |
| --- | --- | --- | --- | --- | --- | --- |
| Class | SVC Precision | SVC Recall | SVC F1-Score | QSVC Precision | QSVC Recall | QSVC F1-Score |
| Reaching for needle with right hand | 0.99 | 1 | 1 | 1 | 0.99 | 1 |
| Positioning needle | 0.98 | 1 | 0.99 | 1 | 1 | 1 |
| Pushing needle through tissue | 0.97 | 0.96 | 0.96 | 0.99 | 1 | 0.99 |
| Transferring needle from left to right | 0.90 | 0.96 | 0.92 | 0.98 | 0.98 | 0.98 |
| Pulling suture with left hand | 0.89 | 0.94 | 0.91 | 1 | 0.99 | 0.99 |
| Dropping suture at end and moving to end points | 1 | 0.95 | 0.98 | 1 | 0.99 | 1 |
| **Accuracy: Classical (0.96), QSVC (0.99)**<br>**Macro Avg: Classical (Precision: 0.95, Recall: 0.97, F1: 0.96), QSVC (Precision: 0.99, Recall: 0.99, F1: 0.99)** |  |  |  |  |  |  |

Its accuracy, precision, recall, and F1-scores for all the gesture classes were better than the Support Vector Classifier (SVC) model. As in the classical model, the QSVC also showed good classification performance for each gesture with almost all gesture classes achieving 100% accuracy. The results of the quantum model also support the application of the quantum model in the recognition of complex surgical tasks, and the accuracy of the quantum model is equivalent to that of the classical model.

In figure 3, the correlation between the test accuracy and the cross-validation scores for both the support vector and quantum support vector was very high but the QSVC seems to learn faster as data increases as compared to the SVC though the SVC performs well as the data increases, this shows that the model is not underfitting because the performance of the model is almost the same for the different validation sets. The decreasing validation loss shows that the model is generalizing better as it is exposed to more data.

**Figure 3:**
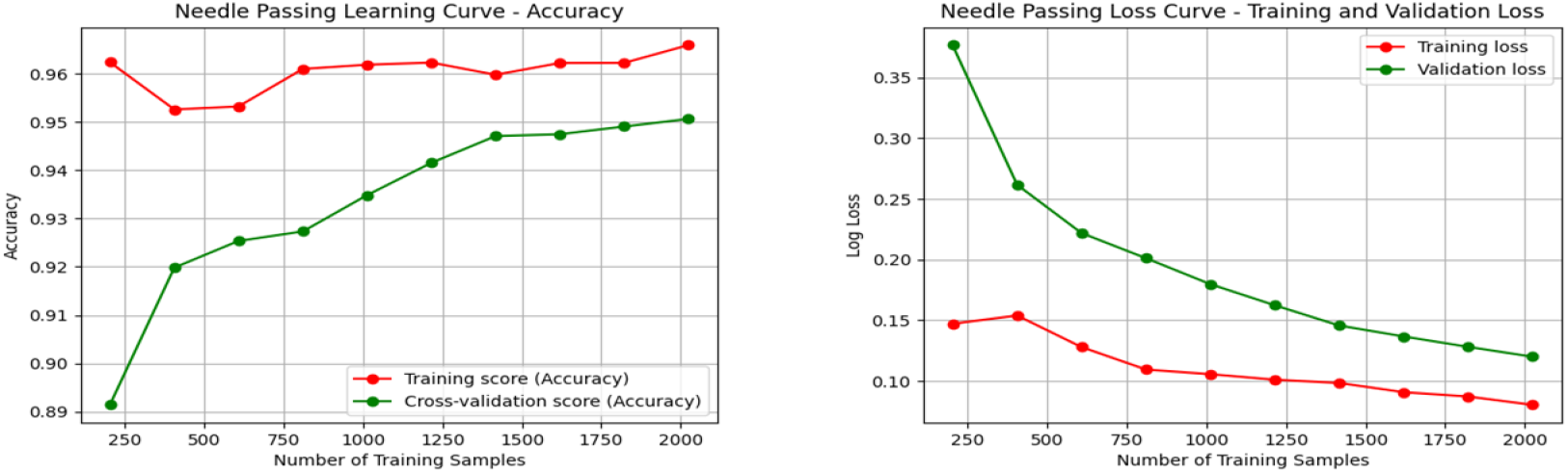
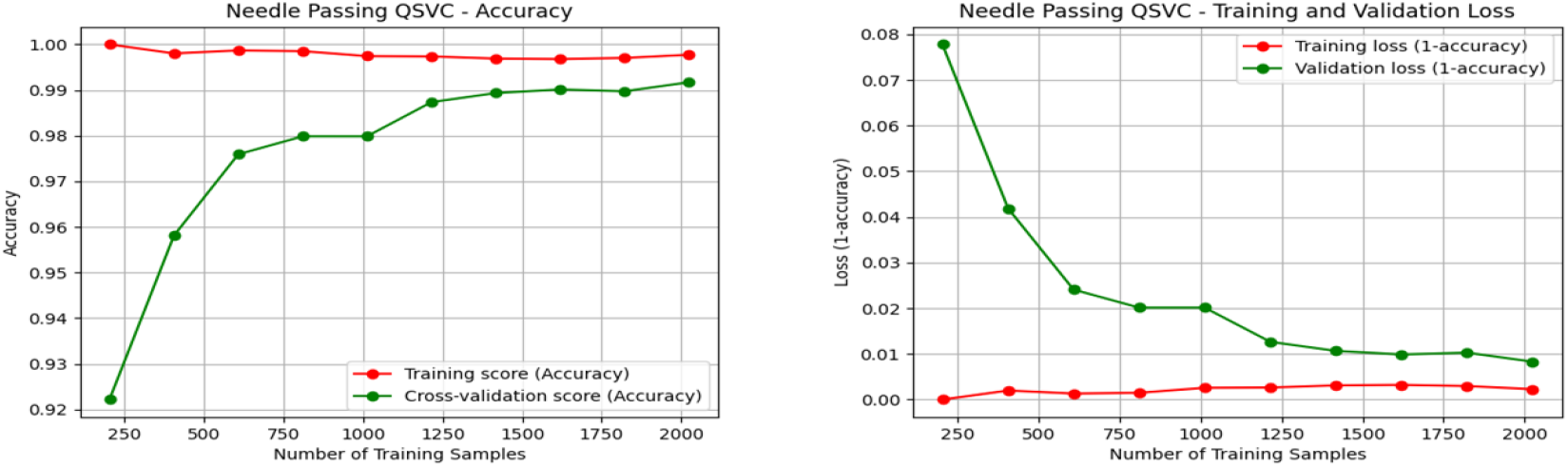
Needle passing Training Accuracy and validation loss

**Figure 4:**
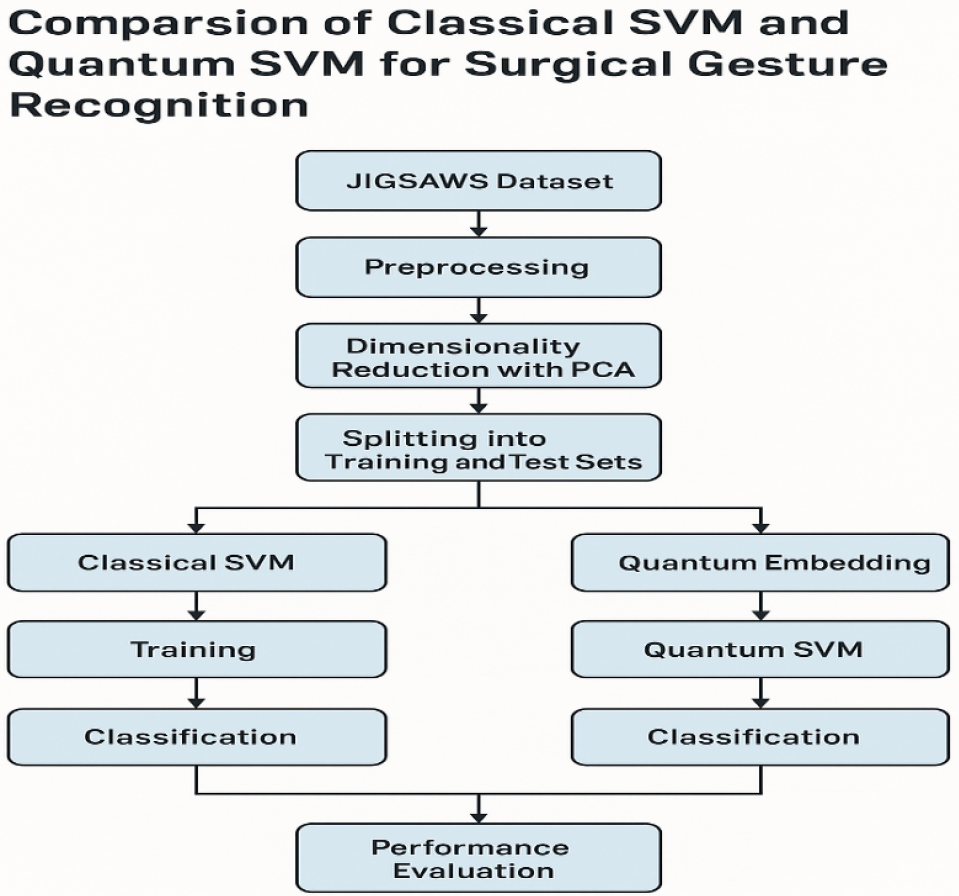
Model Development process

## Discussion

The study only considers QSVC as the quantum model, which limits the assessment of quantum machine learning’s possibilities. Further investigation of other quantum algorithms like Quantum Neural Networks (QNN) or Quantum k-Nearest Neighbors (QkNN) may help to get a better picture of quantum machine learning in the context of surgical gesture recognition. Narrow Focus on the JIGSAWS Dataset: The JIGSAWS dataset only includes three surgical tasks: suturing, knot tying, and needle passing. This limited scope means the study cannot fully assess the models’ performance across a wider variety of surgical gestures. Broader datasets or more complex tasks would provide better insight into the generalizability of the models. The scalability of the QSVC model, especially in larger datasets or more complex surgical procedures, is not addressed. As quantum computing’s ability to handle larger datasets is often cited as an advantage, testing the models on more expansive datasets would provide clearer insights into the future scalability of QML models.

The classical SVM model showed convergence as suggested by the flattening of the loss function and cross validation metrics. The last training iterations demonstrated that the classification accuracy and loss values did not change much, which means that the optimal hyperplane was reached. QSVM model also showed that the optimization was achieved by the stabilization of cost function during the training phase. The updates in the parameters became less frequent in the later training phases, and the performance measures stabilized, which showed that the quantum circuit was able to learn the decision boundary for the surgical tasks. The QSVC performed better than the SVM giving quantum the potential to transform healthcare. The results attained from this study for the SVM is comparable to the study conducted by (Lukács et al., 2023) which achieved an accuracy of 96% which is the same as the results achieved for this study.

### Conclusion and Recommendation

This research work aimed at the surgical procedure recognition and comparing the classical SVM and QSVM for the classification based on kinematic data from JIGSAWS. QSVM had slight enhancements of accuracy in more intricate tasks such as knot-tying which is an indication of quantum computing. However, when the given problem is mapped into a feature space using a Kernel, present quantum hardware limitations make, classical SVM more efficient.

In further research, more of the existing quantum algorithms should be investigated, more comparisons made on computational complexity between both classical and quantum counterparts, as well as application on real-world data. However, further improvements of quantum models for real-time surgical applications, as well as the incorporation of hybrid methods, may improve both efficiency and applicability.

### Future Works

One area for further research is to create a mobile system application that utilizes the identified surgical procedure recognition models for empirical use for real-time or post-operative feedback. A benefit of this application is that it would provide feedback about the surgeons’ performance real-time or after a surgical operation that would enable them improve their surgical skills. Furthermore, it can be incorporated into training applications based on both classical and quantum approaches for analyzing surgeon’s movements and enhancing the training process, as well as minimizing mistakes in practice. This research specifically contributes to achievement of the Sustainable Development Goals 2030 by ensuring good health and well-being (SDG 3).

## Data Availability

The dataset used in this study is the JHU-ISI Gesture and Skill Assessment Working Set (JIGSAWS), which was developed through a collaboration between Johns Hopkins University and Intuitive Surgical Inc. The dataset is publicly available for research purposes upon request and approval from the dataset providers. Access can be obtained by submitting a request through the official project website: https://cirl.lcsr.jhu.edu/research/hmm/datasets/jigsaws_release/

## Source

https://cirl.lcsr.jhu.edu/research/hmm/datasets/jigsaws_release/

## REFERENCES

Cerezo, M., et al. (2022). Challenges and opportunities in quantum machine learning. Nature Computational Science, 2(9), 567–576.

De Maio, V., et al. (2024). Training Computer Scientists for the Challenges of Hybrid Quantum-Classical Computing. arXiv preprint 2403.00885.

Eckhoff, J. A., et al. (2023). TEsoNet: knowledge transfer in surgical phase recognition from laparoscopic sleeve gastrectomy to the laparoscopic part of Ivor-Lewis esophagectomy. Surg Endosc, 37(5), 4040–4053. Retrieved from https://www.ncbi.nlm.nih.gov/pubmed/36932188. doi:10.1007/s00464-023-09971-2

Elgezua, I., et al. (2013). Survey on current state-of-the-art in needle insertion robots: Open challenges for application in real surgery. Procedia CIrP, 5, 94–99.

Fard, M. J., et al. (2016). Toward personalized training and skill assessment in robotic minimally invasive surgery. arXiv preprint 1610.07245.

Farivar, B. S., et al. (2015). General surgery residents’ perception of robot-assisted procedures during surgical training. Journal of surgical education, 72(2), 235–242.

Flöther, F. F. (2023). The state of quantum computing applications in health and medicine. Research Directions: Quantum Technologies, 1, e10.

Gao, X., et al. (2020). Automatic gesture recognition in robot-assisted surgery with reinforcement learning and tree search. Paper presented at the 2020 IEEE international conference on robotics and automation (ICRA).

Hamdi, A., et al. (2021). MARL: multimodal attentional representation learning for disease prediction. Paper presented at the International Conference on Computer Vision Systems.

Hassanzadeh, P. (2020). Towards the quantum-enabled technologies for development of drugs or delivery systems. Journal of controlled release, 324, 260–279.

Huo, C.-M., et al. (2017). Tongue shape classification integrating image preprocessing and convolution neural network. Paper presented at the 2017 2nd Asia-Pacific Conference on Intelligent Robot Systems (ACIRS).

Innan, N., et al. (2023). Enhancing quantum support vector machines through variational kernel training. Quantum Information Processing, 22(10), 374.

Ismail Fawaz, H., et al. (2018). Evaluating surgical skills from kinematic data using convolutional neural networks. Paper presented at the Medical Image Computing and Computer Assisted Intervention–MICCAI 2018: 21st International Conference, Granada, Spain, September 16-20, 2018, Proceedings, Part IV 11.

Jackson, R. C., et al. (2016). Needle-tissue interaction force state estimation for robotic surgical suturing. Paper presented at the 2016 IEEE/RSJ International Conference on Intelligent Robots and Systems (IROS).

Jain, S., et al. (2022). General surgery: requirements, rationale, and robust results. The Surgery Journal, 8(04), e342–e346.

Khan, M. S., et al. (2003). Use of suturing as a measure of technical competence. Annals of plastic surgery, 50(3), 304–309.

Kulkarni, A. D. Challenges of Post-Surgery Recovery and Rehabilitation.

Lin, H. C., et al. (2005). Automatic detection and segmentation of robot-assisted surgical motions. Paper presented at the International conference on medical image computing and computer-assisted intervention.

Lukács, E., et al. (2023). Enhancing Skill Assessment of Autonomous Robot-Assisted Minimally Invasive Surgery: A Comprehensive Analysis of Global and Gesture-Level Techniques applied on the JIGSAWS Dataset. Acta Polytechnica Hungarica, 20(8).

Maniscalco, S., et al. (2022). Quantum network medicine: rethinking medicine with network science and quantum algorithms. arXiv preprint 2206.12405.

Mathur, N., et al. (2021). Medical image classification via quantum neural networks. arXiv preprint 2109.01831.

Matic, A., et al. (2022). Quantum-classical convolutional neural networks in radiological image classification. Paper presented at the 2022 IEEE International Conference on Quantum Computing and Engineering (QCE).

McKnight, R. R., et al. (2020). Virtual reality and augmented reality—translating surgical training into surgical technique. Current reviews in musculoskeletal medicine, 13, 663–674.

Mohan, A., et al. (2013). Operating efficiency: a six-month review. British Journal of Healthcare Management, 19(8), 400–405.

Nasser, D. N. l. (2018). Augmented reality in education learning and training. Paper presented at the 2018 JCCO Joint International Conference on ICT in Education and Training, International Conference on Computing in Arabic, and International Conference on Geocomputing (JCCO: TICET-ICCA-GECO).

Qian, C., et al. (2025). Deep reinforcement learning in surgical robotics: Enhancing the automation level. Handbook of Robotic Surgery, 89–102.

Qin, Y., et al. (2020). Temporal segmentation of surgical sub-tasks through deep learning with multiple data sources. Paper presented at the 2020 IEEE International Conference on Robotics and Automation (ICRA).

Reddy, K., et al. (2023). Advancements in Robotic Surgery: A Comprehensive Overview of Current Utilizations and Upcoming Frontiers. Cureus, 15(12).

Riskin, D. J., et al. (2006). Innovation in surgery: a historical perspective. Annals of surgery, 244(5), 686–693.

Sarikaya, D., et al. (2020). Towards generalizable surgical activity recognition using spatial temporal graph convolutional networks. arXiv preprint 2001.03728.

Sarker, S. K., et al. (2005). Errors in surgery. International Journal of Surgery, 3(1), 75–81.

Shen, Z., et al. (2018). A novel clinical-simulated suture education for basic surgical skill: suture on the biological tissue fixed on standardized patient evaluated with objective structured assessment of technical skill (OSATS) tools. Journal of Investigative Surgery, 31(4), 333–339.

Solenov, D., et al. (2018). The potential of quantum computing and machine learning to advance clinical research and change the practice of medicine. Missouri medicine, 115(5), 463.

Srikanth, P., et al. (2022). Secure quantum computing for healthcare sector: A short analysis. arXiv preprint 2211.10027.

Tarassoli, S. P. (2019). Artificial intelligence, regenerative surgery, robotics? What is realistic for the future of surgery? Annals of Medicine and Surgery, 41, 53–55.

